# Machine Learning Can Predict Deaths in Patients with Diverticulitis During their Hospital Stay

**DOI:** 10.1101/2020.02.04.20020222

**Authors:** Fahad Shabbir Ahmed, Raza-Ul-Mustafa, Liaqat Ali, Imad-ud-Deen, Tahir Hameed, Asad Ikram, Syed Ahmad Chan Bukhari

## Abstract

**Introduction:** Diverticulitis is the inflammation and/or infection of small pouches known as diverticula that develop along the walls of the intestines. Patients with diverticulitis are at risk of mortality as high as 17% with abscess formation and 45% with secondary perforation, especially patients that get admitted to the inpatient services are at risk of complications including mortality. We developed a deep neural networks (DNN) based machine learning framework that could predict premature death in patients that are admitted with diverticulitis using electronic health records (EHR) to calculate the statistically significant risk factors first and then to apply deep neural network.

**Methods:** Our proposed framework (Deep FLAIM) is a two-phase hybrid works framework. In the first phase, we used National In-patient Sample 2014 dataset to extract patients with diverticulitis patients with and without hemorrhage with the ICD-9 codes 562.11 and 562.13 respectively and analyzed these patients for different risk factors for statistical significance with univariate and multivariate analyses to generate hazard ratios, to rank the diverticulitis associated risk factors. In the second phase, we applied deep neural network model to predict death. Additionally, we have compared the performance of our proposed system by using the popular machine learning models such as DNN and Logistic Regression (LR).

**Results:** A total of 128,258 patients were used, we tested 64 different variables for using univariate and multivariate (age, gender and ethnicity) cox-regression for significance only 16 factors were statistically significant for both univariate and multivariate analysis. The mortality prediction for our DNN out-performed the conventional machine learning (logistic regression) in terms of AUC (0.977 vs 0.904), training accuracy (0.931 vs 0.900), testing accuracy (0.930 vs 0.910), sensitivity (90% vs 88%) and specificity (95% vs 93%).

**Conclusion:** Our Deep FLAIM Framework can predict mortality in patients admitted to the hospital with diverticulitis with high accuracy. The proposed framework can be expanded to predict premature death for other disease.

## Introduction

Diverticulitis of the colon is the inflammation of the colon segment that has developed outpouching that most commonly occurs in the sigmoid colon. this occurs secondary to the mucosal and submucosal layers of that segment have herniated in the muscular layer of the bowel wall. This disrupts the normal bowel function of the affected bowel segment and cases stasis of fecal matter in that bowel segment leading to accumulation of fecal matter and subsequent bacterial growth and inflammation. in the United States diverticulitis is the eight most common gastrointestinal diagnosis in the outpatient setting with an estimated 2.7 million annual health care visits. This carries an approximate burden of 2.2 billion dollars (1)

Mortality predication using machine learning has now gained momentum in the past few years particularly using deep neural network (2, 3). The ability of DNN to recognize complex patterns helps it to learn about biological risk factors and predict outcomes based on that learning (4, 5). Simply, machine learning is a branch of computer science that deals with the estimation of an outcome based on past experiences using a computer-generated algorithm (6). We set out to create a prediction system that can predict mortality with high accuracy in patients that were admitted to hospital patients with the primary diagnosis of diverticulitis We used FLAIM (machine learning framework) previously to predict mortality in ICU patients (7) and a comparative Logistic regression (LR) model (8). We hypothesized that the DNN would outperform the LR model.

## METHODS

### Dataset and patient selection

We obtained a publicly accessible data-set National Inpatient Sample (NIS) for the year 2014. This database is part of Health-care Cost and Utilization Project (HCUP). NIS is the largest database available with all-payer inpatient health care in the United States. Contains about 7 million admissions per year approximately. All this data is de-identified. All the database tables were aggregated using “key-nis” variable. Case selection was done using the ICD-9 codes for diverticulitis without hemorrhage (562.11) and diverticulitis with hemorrhage (562.13).

### Data analysis

Patient demographics were calculated and are presented in table 1. A Univariate and Multivariate Cox regression analysis was done using SPSS 24.0 was done to evaluate the role of commodities and other parameters to assess the mortality risk associated with this risk factors. The data-set was sorted and binarized to be used for machine learning.

**Table 1.**
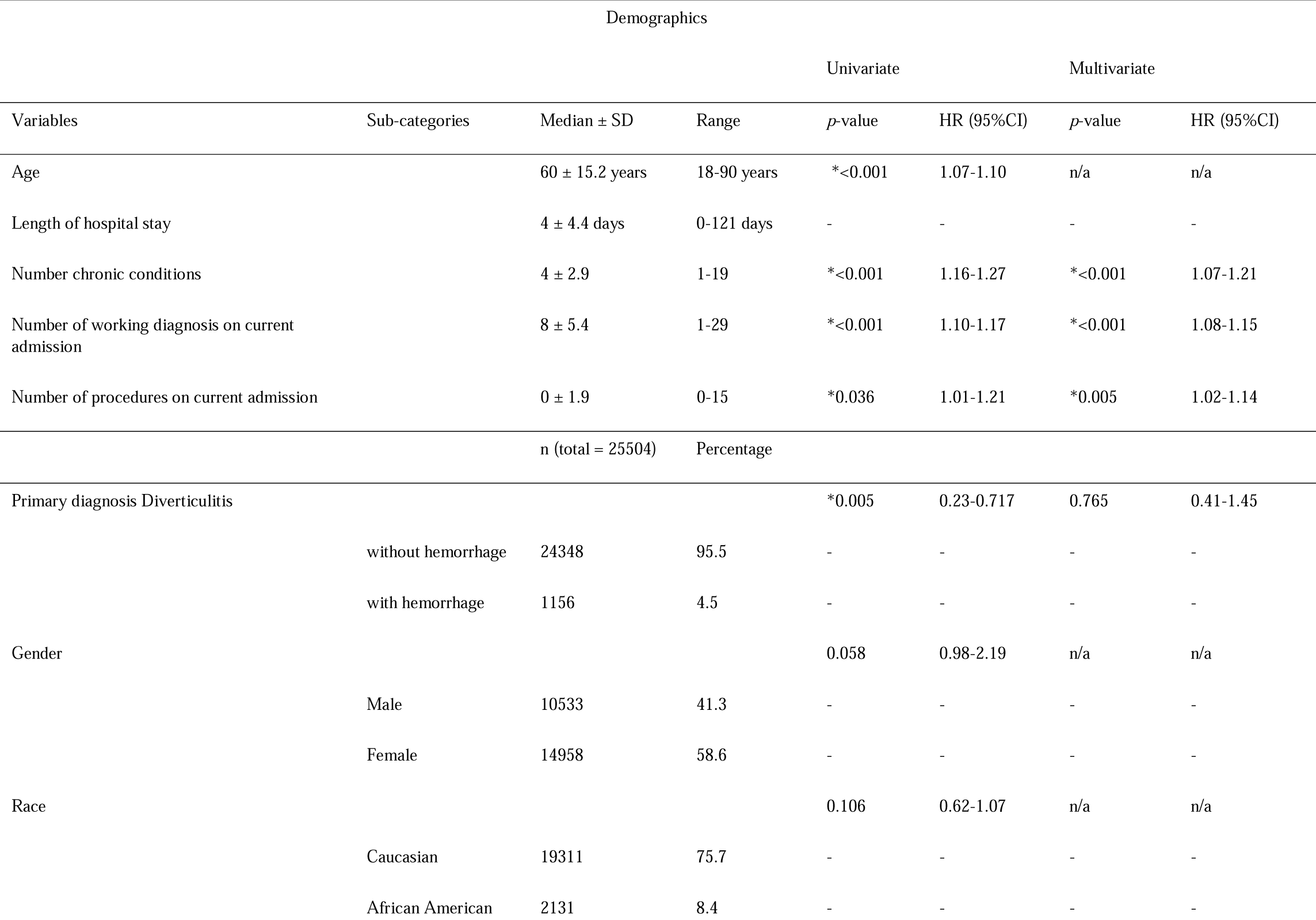
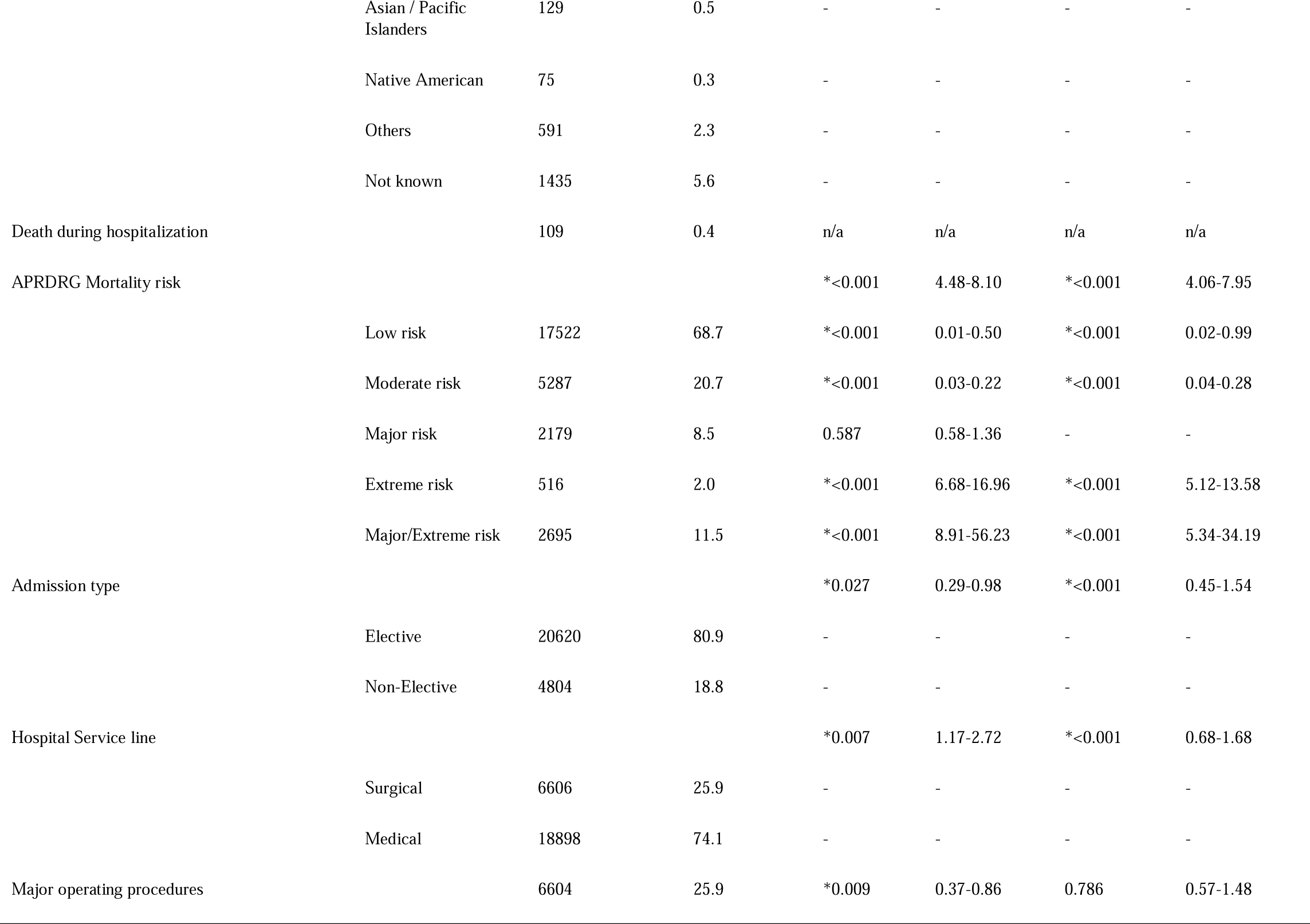
Demographics

### Machine Learning Methods

Logistic Regression (LR) is the appropriate regression analysis to conduct when the dependent binary variable. To all the regression analyses, logistic regression is a predictive analysis. Moreover, it is used to describe data and to explain the relationship between one dependent binary variable and one or more nominal, ordinal, interval or ratio level independent variables. Later, we used NN for the reason it does not impose any restrictions on the input variables (like how they should be distributed). Additionally, many studies have shown that NN can better model i.e. data with high volatility and non-constant variance, given its ability to learn hidden relationships in the data without imposing any fixed relationships in the data. We used Keras a python library that has been used for efficient numerical computations It is high level Application Programming Interface (API) for deep learning models to build and train. Moreover, it wraps Theano and TensorFlow. For fast numerical computational purposes Theano is an efficient library. The parameters we have used for the training and testing is listed in Table 1. On hidden layers we used rectifier and for the output layer we used sigmoid activation function. Finally, the network uses the efficient Adam gradient descent optimization algorithm with a logarithmic loss function, which is called binary-cross entropy in Keras. The performance of the model on the train and test sets recorded during training graphed using a line plot, one for each of the loss and the classification accuracy as shown in Figure 1.

**Figure 1.**
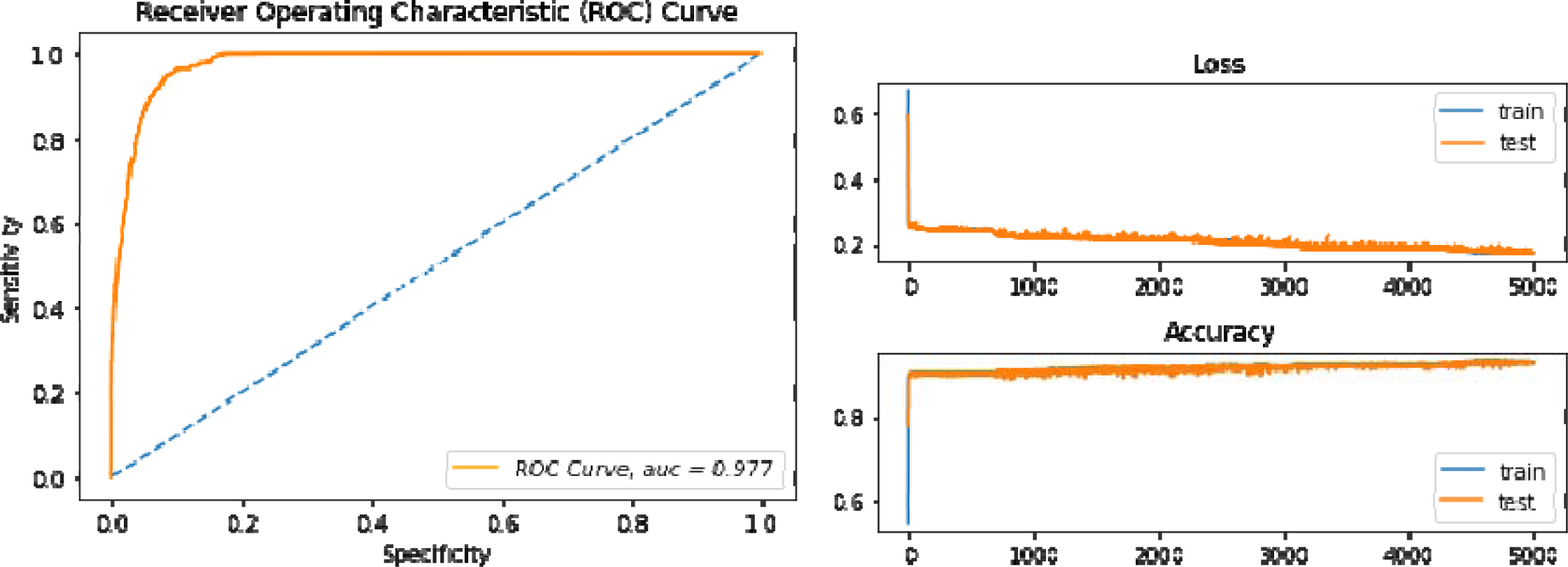
Receiver -Operator Curve for FLAIM-DNN-4RS and the loss and accuracy curve during training.

To further investigate the performance of the proposed method, another evaluation metric i.e., Receiver Operating Characteristic (ROC) chart is used. The ROC chart is a plot of true positive rate (TPR) versus false positive rate (FPR) for various thresholds. An ROC chart with more area under the curve (AUC) is considered best. An ideal ROC chart has AUC=100, such chart means that the model is capable of performing with 100 % sensitivity and 100 % specificity.

## RESULTS

From a total of 3.8 million patients 22504 adult patients (age more than 18 years) were extracted with the primary diagnosis of diverticulitis that were with the median age of 60 years, with most patients being women (58.6%) with mostly being Caucasian (75.7%). the median stay in the hospital was 4 / 4.4 days. These patients had a median stay of 14.5 16.2 days of hospital stay and 4.2 10.7 days of intensive care stay. Emergency admission comprised of most admission making 76.7% of all patients that were admitted to the ICU while most admission came through referrals around 490 patients (48%). Mortality among these patients was 16.8% (173), while 441 (43.2%) were transferred out to different health facilities and 407 (40%) patients were discharged home. All of this can be seen in Table 1.

We conducted a univariate and multivariate analysis of a number of different risk factors and the risk factors that were statistically significant for mortality in the multivariate analysis were: higher number chronic conditions, higher number of working diagnosis on admission, higher number of procedures in the current admission. APRDRG high risk and major risk, non-elective admission, Chronic Heart Failure, Chronic Lung Disease, Coagulopathy, Fluid and electrolyte disorder, Renal Failure, Valvular disease and Weight loss. Other statistically significant risk factors included median house hold income (Upper 50th vs Lower 50th). More detailed statistics can be found on in Tables 2. And 3.

**Table 2.**
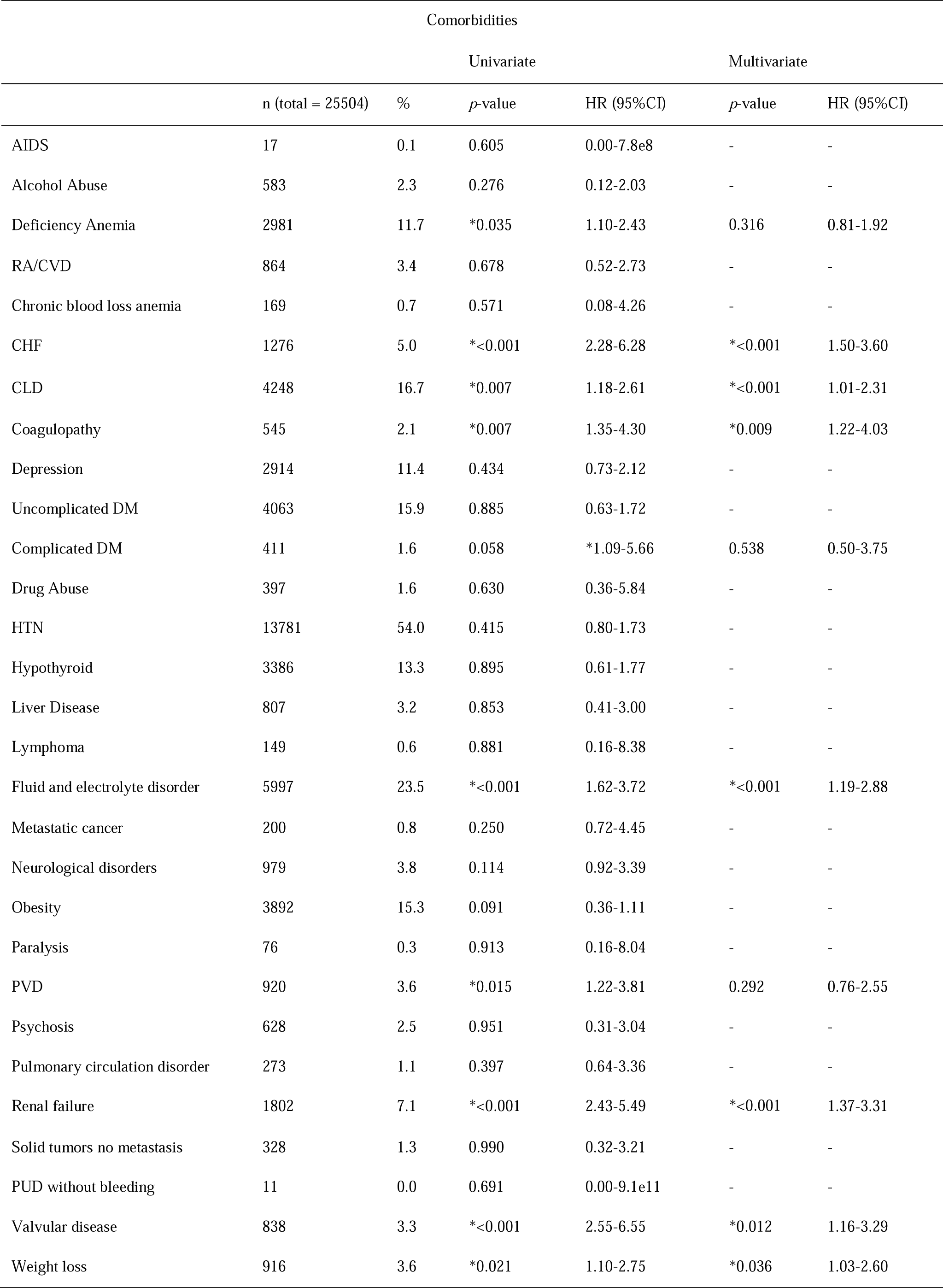
Comorbidities

**Table 3.**
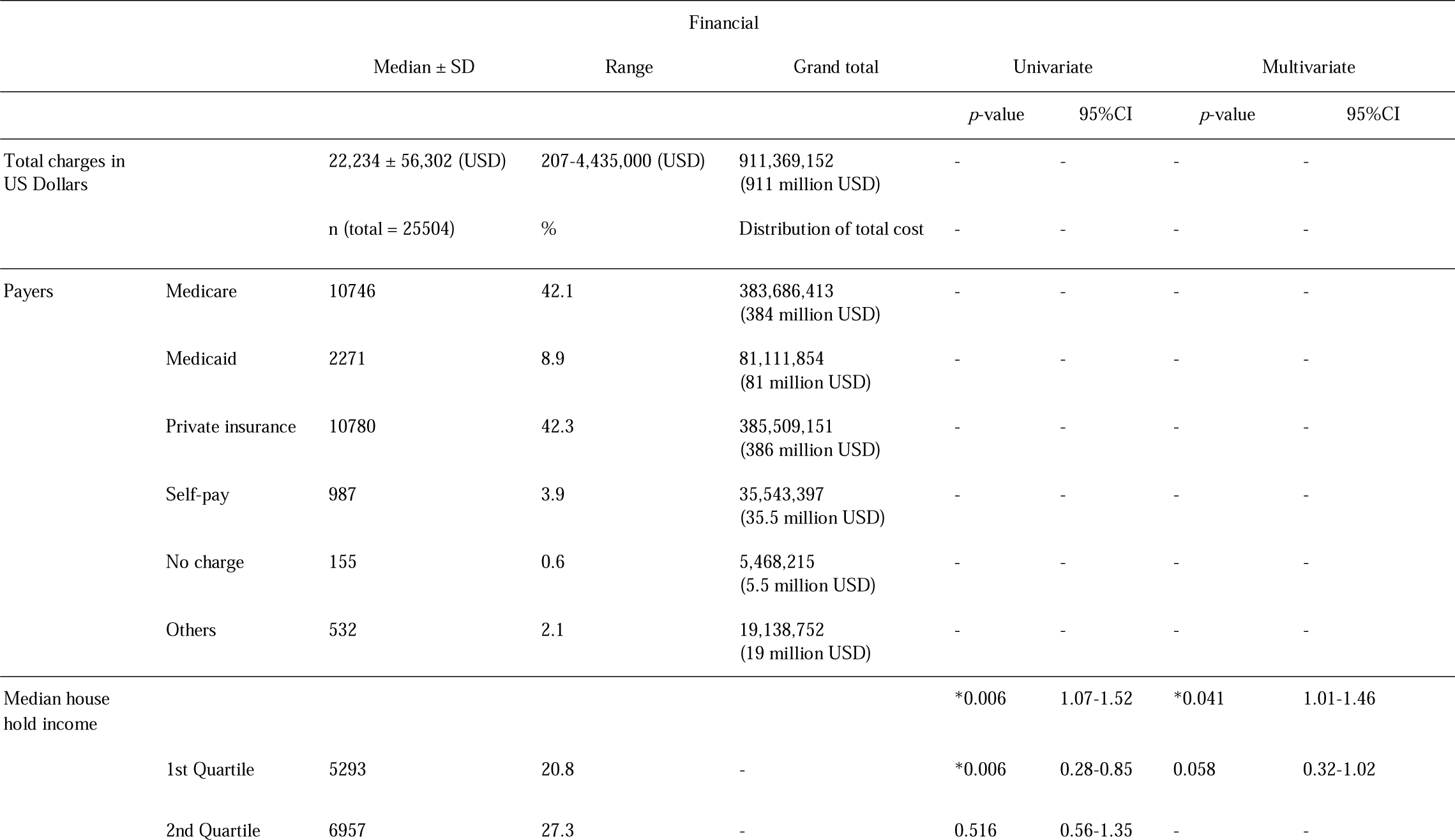
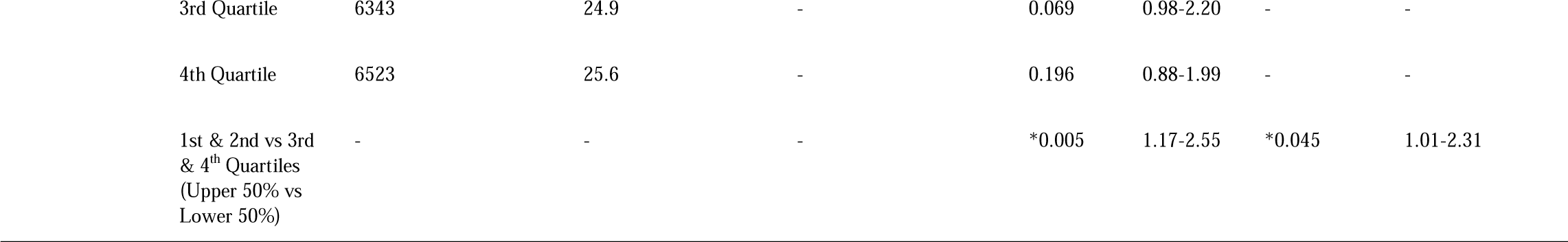

The FLAIM-DNN-4RS showed superior performance compared to the logistic regression model in terms of Training set accuracy (93.1% vs 90.0%), Test set accuracy (93.0% vs 0.91%), Sensitivity specificity and area under the receiver-operator curve which are represented by Table 4 and Figure 1. Figure 1 also shows the loss and accuracy during training for FLAIM-DNN-4RS. Table 5 shows the details of the FLAIM-DNN-4RS model.

**Table 4.**
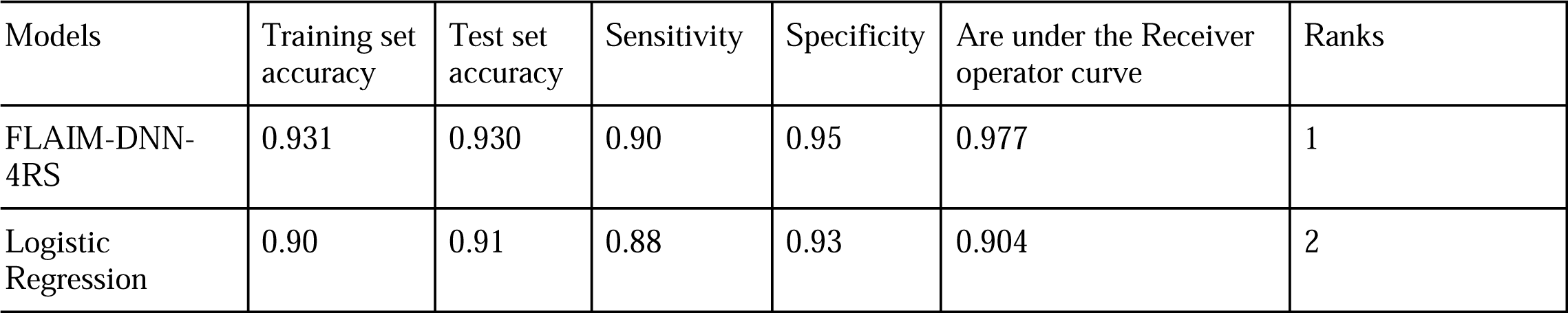
Machine Learning Results for FLAIM-DNN-4RS and the Logistic regression model

**Table 5:**
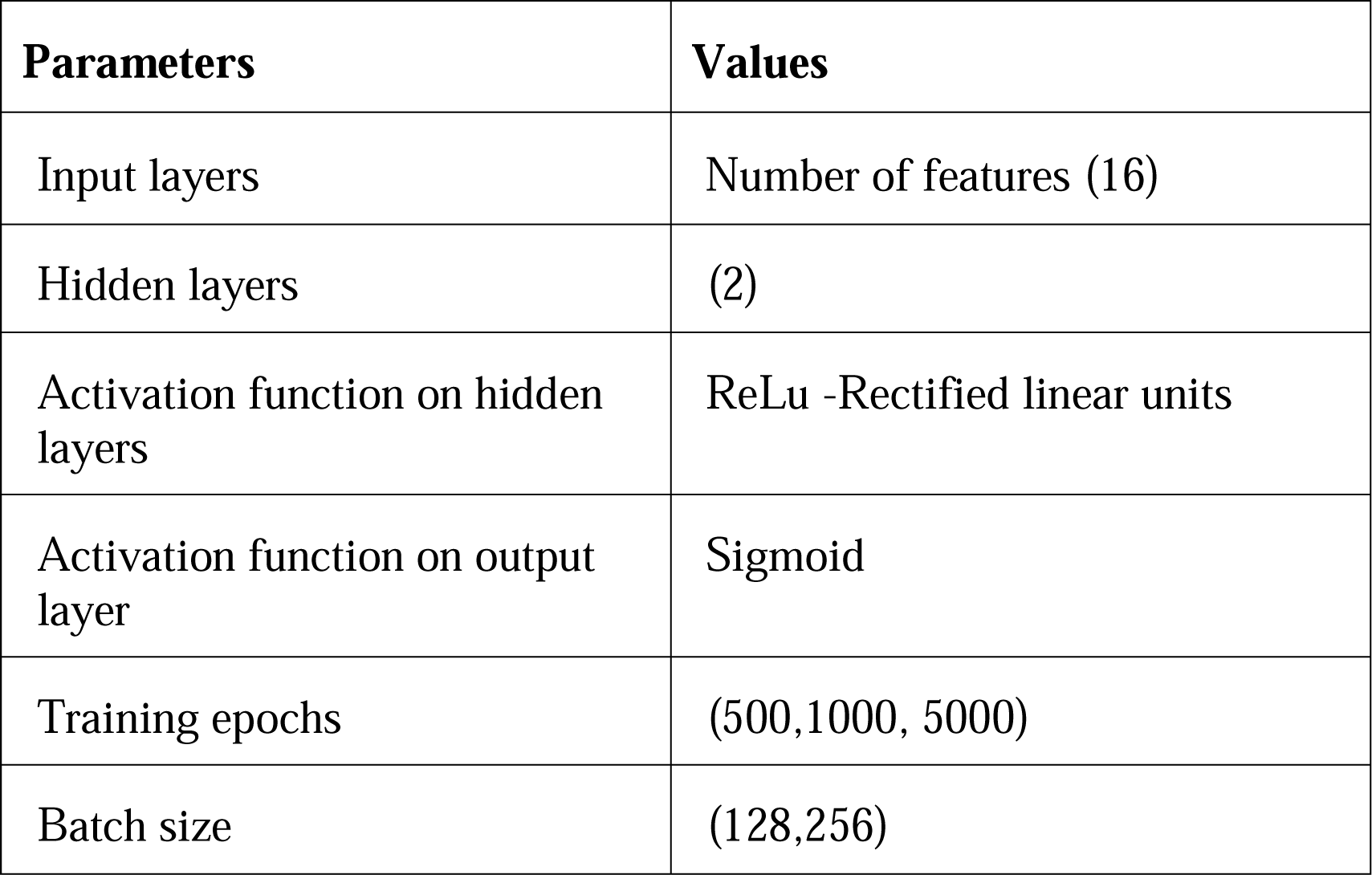
Parameters used during training and testing of proposed method

## DISCUSSION

The incidence of diverticular disease has dramatically increased over the years. Current data suggest that up to 50% of individuals older than 60 years of age suffer from diverticular disease and its complications resulting in a sharp rise of hospitalizations (9).

The current study confirmed high mortality risk in patients of more than 60 years of age (p<0.001). It was also seen that the number of chronic conditions also affected the overall mortality of these patients(p<0.05) admitted with acute diverticulitis significantly, with chronic liver disease, fluid and electrolyte imbalance, Renal failure, weight loss and valvular heart disease (p<0.001). We also looked at the clinical risk factors such as chronic NSAID use, Bisphosphonate use, tobacco use, steroid use and antibiotic use which did not affect mortality.

We compared the median household income of the patients and found an increased risk of mortality in the lower 50% of the population (3rd and 4th quartiles (p<0.005)). This might be due to better healthcare access, insurance; and possibly education, dietary habits and a healthier lifestyle in patients having household income with in the 3rd and 4th quartile range. This was in contrast to a study conducted in Sweden which showed patients with middle to high household income had a higher risk of diverticular diseases when compared with low household income (10).

Through the application of machine learning methods to the various data present at the time of the new diagnosis of diverticulitis, disease mortality can be moderately predicted (11). The Machine Learning algorithms used in this analysis can provide mortality prediction based upon the values of blood biomarkers at the time of admission and at different time intervals. Disease prediction though this model can help in the prognosis of the disease leading to better outcomes. It can also be used for risk stratification or help providers in clinical decision-making. For diverticulitis, this type of model can also be used to aid in decisions made by physicians and patients. Though future reiteration of this model must be validated using independent samples for a different dataset. (12).

## CONCLUSION

FLAIM-DNN-4RS is a robust model that can predict mortality in diverticulitis patient with very high accuracy we need to further validate this model.

## Data Availability

This is a publically available database.

## Notes

### Competing Interest Statement

The authors have declared no competing interest.

### Funding Statement

None

